# Structural alterations in a rumination-related network in patients with major depressive disorder

**DOI:** 10.1101/2023.11.10.23298406

**Authors:** Paul Z. Cheng, Hsin-Chien Lee, Timothy J. Lane, Tzu-Yu Hsu, Niall W. Duncan

## Abstract

Rumination is a common symptom in major depressive disorder (MDD) and has been linked to risk for that disorder, its prognosis, and relapse likelihood. Previous work has linked individual differences in rumination to structural properties in a variety of brain regions. Some of these regions, such as the dorsolateral prefrontal cortex (dlPFC), have also been highlighted as being altered in MDD, suggesting a connection between structural changes and ruminative symptoms. Although informative, such localised relations have some limitations in the context of a network view of the brain. To further investigate rumination-related structural changes in depression and to situate these within potential functional networks, we acquired structural data from patients with MDD (n = 32) and controls (n = 69). Analysis of cortical grey-matter identified group differences in the dlPFC that were, however, not related to rumination. Instead, rumination was correlated with grey-matter properties in the right precuneus. Using normative functional connectivity analysis on a large independent sample, we show that these two regions are interconnected. It was further shown that the rumination-related precuneus region is highly connected with networks associated with processes such as executive function, autobiographical memory, and visual perception. Notably, each of these processes has been connected to rumination. These results suggest that rumination in depression may be linked to focal structural changes that disrupt a distributed functional network.

## Introduction

Major depression (MDD) is a disorder with varied symptomatology that has been associated with structural alterations in different brain regions. In particular, grey-matter volume in several brain regions have been found to be reduced in patients with MDD, as compared to healthy individuals. These include the dorsomedial prefrontal cortex (dmPFC), posterior cingulate cortex (PCC), anterior insula, bilateral basal ganglia, and parahippocampal gyrus (Arnone et al., 2016; Bora et al., 2012; Denson et al., 2009; Espinoza Oyarce et al., 2020; Hooker et al., 2010; Klok et al., 2019; Koolschijn et al., 2009; Kross et al., 2009; Lemogne et al., 2009; Machino et al., 2014; Putnam and McSweeney, 2008; Schmaal et al., 2017; Wang et al., 2015). Although these MRI studies of brain structure have provided evidence for anatomical changes in MDD generally, questions remain regarding the relationships between alterations in particular brain regions and specific symptoms of the disorder.

One common symptom of MDD is depressive rumination, which is characterised by repetitive focus upon negative thoughts, feelings, and events in one’s life (Nolen-Hoeksema, 1991). Clinically, rumination has been found to predict the onset (Abela and Hankin, 2011) and severity (Nolen-Hoeksema et al., 2008) of depressive episodes, the likelihood of relapse in those recovering from the condition (Michalak et al., 2011; Spinhoven et al., 2018), and likelihood for healthy individuals developing depression (Spasojević and Alloy, 2001). Given this connection to clinical progression, investigating the structural changes in the brain underlying rumination is likely to be of importance for understanding MDD.

Studies in healthy participants have identified correlations between rumination and grey-matter volumes. For example, a negative correlation between rumination and grey-matter volume has been observed in the bilateral inferior frontal gyrus (IFG), left anterior cingulate cortex, and bilateral mid-cingulate cortex (Kühn et al., 2012). The overall pattern of results has been mixed, however, with other studies reporting a positive correlation between rumination and regional grey-matter volume in the dorsolateral prefrontal cortex (dlPFC) and parahippocampal gyrus (Wang et al., 2015). Taking the brooding and reflective pondering subscales of rumination questionnaires in particular, both positive and negative correlations, respectively, have been reported with the left dlPFC and dmPFC (Sin et al., 2018). Some variety in outcomes can also be observed in studies looking at rumination in patients with MDD. Here, rumination has been reported to be negatively correlated with grey-matter volume in the dlPFC (Wang et al., 2015) and positively correlated with grey-matter volume in the right superior temporal gyrus (Machino et al., 2014). Taken together, these results point to there being a link between grey-matter changes in the brain and rumination but leave open questions about the specific nature of any such connection.

The role of individual brain regions can be more fully understood in the context of their involvement in the dynamic networks that span the brain (Javaheripour et al., 2023; Seguin et al., 2023). Structural changes in one region have been shown to influence functional activity in diverse other regions, with these changes then being related to behavioural alterations (Fox, 2018). For example, particular functional networks centred around structural lesions have been connected to mental phenomena such as manic symptoms (Cotovio et al., 2022), mind wandering (Philippi et al., 2021), and executive function (Hwang et al., 2020). Given the mixed results to date relating brain structure to rumination in MDD, it may be the case that the specific behavioural changes observed are the result of dispersed network effects in the disorder (Drysdale et al., 2017; Siddiqi et al., 2021). Potential connections of this type between structural changes and functional networks have begun to be studied, but such relationships remain to be more fully explored. In particular, it is not clear whether rumination symptoms in depression may be connected to particular focal structural changes or if they are linked to more dispersed functionally connected sets of regions.

To investigate structural alterations in MDD and their potential link to rumination, we acquired structural MRI data from a set of patients with MDD and healthy controls. This data was then combined with a set of normative functional connectivity data for analysis (Boes et al., 2015). The aims of the study were two-fold: 1) determine whether group difference brain regions and rumination correlated brain regions are overlapping or distinct; and 2) identify the functional networks within which any brain structural properties linked to rumination may be embedded. It was anticipated that structural differences at the group level would be seen in regions such as the dlPFC, dmPFC, and PCC/precuneus. It was further supposed that these structural differences would be correlated with rumination and would be located within a shared functional network.

## Methods

### Participants

One hundred and one participants were recruited. Thirty-two were patients with MDD (mean age = 38.4 ± 13.5 years; 26 female) and the remaining 69 healthy controls (mean age = 33.6 ± 11.6 years; 54 female). Individuals with MDD were recruited from the Taipei Medical University-Shuang Ho Hospital psychiatric day clinic. Diagnosis was made according to DSM-IV-TR criteria. Most individuals with MDD (30 out of 32) were medicated at the time of participation, with an average of 8.1 months (± 14.5) since the onset of their current episode.

Control group participants were recruited from the local population in New Taipei City through referrals and advertisements at Shuang Ho Hospital. They all reported no history of neurological or psychiatric disorders, nor were any using psychoactive medication at the time of participation. Three control group participants did not complete the MRI scanning and so were excluded.

All participants reported no history of neurological disorders or head injury, current use of recreational drugs, metal implants, pregnancy, claustrophobia, or other MRI contraindications. Written informed consent was obtained from all participants and they were compensated financially for their participation. The study was approved by the Taipei Medical University JIRB (N201603080). An independent analysis of MRI data from the same participants has been published previously (Truong et al., 2021).

### Psychological questionnaires

Questionnaires were administered during the scan day. Traditional Chinese translations of each were used. Individuals with MDD had their depression symptom severity assessed with the Montgomery–Åsberg Depression Rating Scale (MADRAS). Rumination was assessed in all participants through the Ruminative Responses Style (RRS) questionnaire (Huang et al., 2015; Treynor et al., 2003). In addition, depressive symptoms were assessed in all participants through the Beck Depression Inventory II (BDI; Beck et al., 1996; Lu, 2002). Participant characteristics were compared between groups through Welch’s t-tests.

### Imaging data acquisition

Neuroimaging data were acquired on a 3 Tesla GE Discovery MR750 scanner (DV24.0 software version) using a body coil for transmission and an eight-channel receive head coil. A fast spoiled gradient echo (FSPGR) sequence was used to obtain whole-brain T1-weighted structural images (176 sagittal slices, TR/TE/flip angle = 8 ms/3 ms/ 12° matrix = 256 x 256 mm^2^; voxel resolution = 1 x 1 x 1 mm^3^). During scanning, participants’ heads were secured with sponge pads and they were instructed to minimise their movements.

### Quality control

Image quality was appraised using MRIQC (Esteban et al., 2017). Three quality metrics - part of Mortamet’s quality index (qi_2); the standard deviation of foreground-to-background energy ratio (summary_bg_stdv); and the overlapping difference of cerebrospinal fluid tissue probability maps between T1 image and ICBM nonlinear-asymmetric 2009c template (summary_tpm_csf) - were selected based on previously reported importance (Esteban et al., 2017; Mortamet et al., 2009; Shehzad et al., 2015). Each metric was compared between patients and controls with Welch’s t-tests.

### Preprocessing

Anatomical images were processed using Freesurfer’s automatic surface reconstructions, resulting in white matter and pial surface segmentation (Dale et al., 1999). The segmentation accuracy was visually checked for each participant. The resulting cortical surface area and thickness maps were downsampled with the "fsaverage4" template (2562 vertices per hemisphere) and smoothed with a 10 mm FWHM Gaussian kernel (Fischl and Dale, 2000).

### Statistical analysis

Grey-matter volumes at each cortical vertex were defined and analysed through a joint analysis of surface area and thickness using a Fisher nonparametric combination method (Winkler et al., 2018) implemented in the FSL library’s PALM tool (Winkler et al., 2014).

Three linear models were applied to the data in line with the three main aims of the study. These were:

1. A comparison of grey-matter volume between patients and controls to replicate prior findings
2. A regression of RRS scores against grey-matter volume to identify areas where these are related, independent of diagnostic status
3. A test of the interaction between RRS scores*grey-matter volume with respect to diagnostic status

All models included the participant’s age and sex, along with the individual’s intracranial volume, as covariates. Statistical significance testing was performed through permutation tests (500 permutations) with a tail approximation acceleration method (Winkler et al., 2014). Multiple comparisons were controlled with threshold-free cluster enhancement (Smith and Nichols, 2009). Bihemispheric comparisons were accounted for with family-wise error rate (FWER) correction across modalities, with statistical significance defined at P_FWE_ < 0.05 (Alberton et al., 2020). Unthresholded effect size images are also presented (Chen et al., 2022).

### Normative functional connectivity

A seed-based functional connectivity analysis was conducted based on the results of rumination-related cortical surface area changes. This was done on 100 unrelated participants from the Human Connectome Project dataset (Van Essen et al., 2013). The purpose of this analysis was to establish a normative connectivity network for rumination-related grey-matter regions in healthy participants. This aims to establish a network baseline state in a large sample with high-quality functional data. The dataset was preprocessed using the HCP functional preprocessing pipeline (Glasser et al., 2013; Smith et al., 2013). In addition, the data were bandpass filtered between 0.01 and 0.1 Hz.

A seed mask was created from the peak vertex in the rumination-related cluster. This vertex was smoothed to the 10 nearest neighbours to create a circular mask and sampled to human connectome fs_LR_32k functional space. Functional connectivity was defined as Pearson’s correlation between the mean seed time series and all other cortical vertices. A minimum effect size of interest of r = 0.1 was used to identify seed-associated networks, with significance defined with a vertex-wise FDR corrected threshold of p < 0.05. Resulting connectivity values were then arranged according to the Yeo seven-network scheme (Yeo and Krienen, 2011). Connectivity with regions of between-group grey-matter volume differences were also assessed by extracting mean values in any such regions and testing against zero through a one-sample t-test.

### Software versions

Original DICOM images were converted to NIFTI format with dcm2niix (v1.0.20181125). Image preprocessing and statistical analyses were conducted with a combination of the following packages: FSL (5.0.11), FreeSurfer (6.0.0), Nipype (1.2.0), Nitime (0.9), PALM (alpha115), MRIQC(22.0.0rc1) and Python (3.6.12). Analyses were conducted on a Linux 11.0 Intel i7-4790 desktop computer.

## Results

### Participant characteristics

Ninety-eight participants were included in the final analysis. Demographic details are presented in Table 1. Participants with MDD reported higher rumination and severity of depression than controls (RRS: t(46.52) = 7.01, p = 8.37*10^-9^; BDI: t(36.82) = 7.21, p = 1.49*10^-8^).

**Table 1:** Participant characteristics.

|  |  | Missing | Total | MDD | Control |
| --- | --- | --- | --- | --- | --- |
| N |  |  | 98 | 32 | 66 |
| Age, mean (SD) |  | 0 | 35.2 (12.4) | 38.4 (13.5) | 33.6 (11.6) |
| Sex, n (%) | Female | 0 | 80 (81.6) | 26 (81.2) | 54 (81.8) |
|  | Male |  | 18 (18.4) | 6 (18.8) | 12 (18.2) |
| Handness, n (%) | Left | 0 | 6 (6.1) | 2 (6.2) | 4 (6.1) |
|  | Right |  | 92 (93.9) | 30 (93.8) | 62 (93.9) |
| MADRAS, mean (SD) |  | 0 | 20.7 (9.7) | 20.7 (9.7) | N/A |
| BDI, mean (SD) |  | 2 | 13.9 (13.3) | 27.2 (13.8) | 7.9 (7.4) |
| RSS, mean (SD) |  | 3 | 48.4 (14.9) | 62.1 (13.2) | 42.3 (11.2) |
| Reflective rumination, mean (SD) |  | 3 | 10.7 (3.6) | 12.3 (3.2) | 10.0 (3.5) |
| Brooding rumination, mean (SD) |  | 3 | 11.3 (3.8) | 14.8 (3.5) | 9.8 (2.8) |

### Neuroimage quality

To ensure differences found in our samples were not driven by image quality, we compared two groups’ image parameters generated from MRIQC. No differences in T1 image quality parameters between the control and MDD participants were seen (qi_2: t(50.96) = −0.77, p = 0.44; summmary_bg_stdv: t(77.45) = 0.58, p = 0.56; tpm_overlap_csf: t(53.59) = −1.99, p = 0.24).

### Group structural differences

Depression-specific structural differences between groups were assessed by comparing volumetric values between groups. The bilateral inferior frontal gyri, extending to the dlPFC in the left hemisphere, were found to have higher cortical volume in individuals with MDD than in the control group, illustrated in Figure 1A. This group difference appears driven by cortical surface area changes, rather than cortical thickness (Supplementary figure 1).

**Figure 1:**
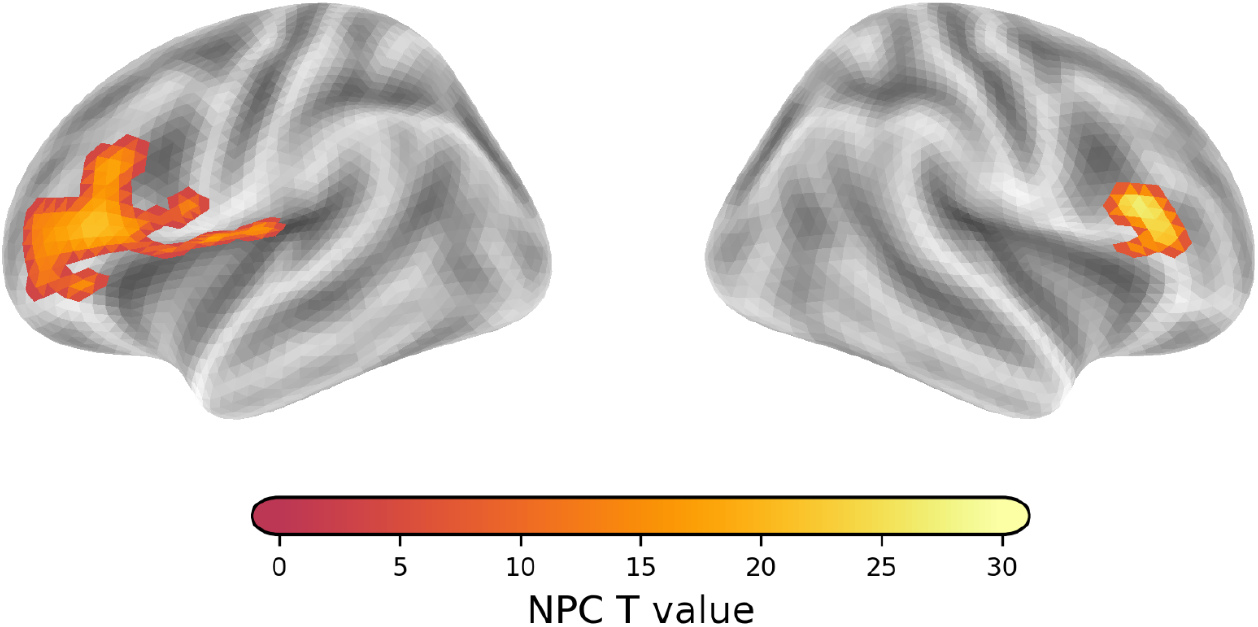
Comparison of grey-matter volume between MDD and control groups. Greater grey-matter volumes were observed in patients with MDD in bilateral dlPFC regions. Statistical comparison was done via NPC Fisher thresholding with P_FWE_< 0.05.

### Rumination-related structural properties

Rumination-related structural effects were assessed by including volumetric values with group and RRS as a regressors of interest. No relationship with the combined cortical volume measure was found. Instead, cortical surface area in the right precuneus region was found to have a significant negative association with rumination across all participants (Figure 2A). In other words, greater reported rumination is related to lower surface area in the right precuneus. This relationship was not found to differ between patients and controls.

**Figure 2:**
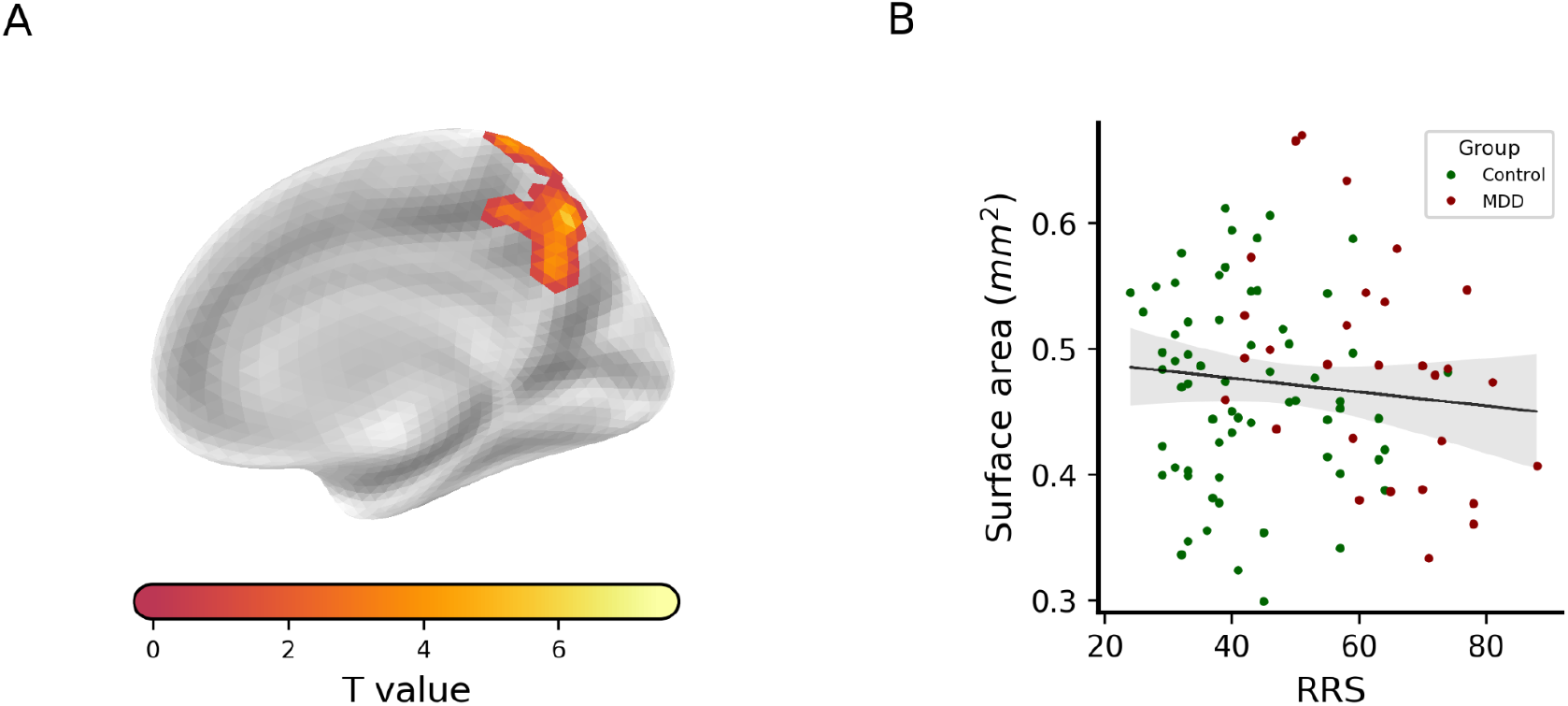
Association between rumination and cortical surface properties. (A) A negative association between the right precuneus region’s surface area and rumination was observed. (B)This association is illustrated through a plot of RRS scores against surface area values from the right precuneus cluster. Note that the association did not differ between MDD and control groups.

### Normative functional connectivity

The region in the right precuneus that was associated with reported rumination was found to be functionally connected to an extensive cortical network (Figure 3A). Subdividing this into the seven canonical networks reported by Yeo et al. (2011), the region was found to be particularly richly connected to the default-mode, fronto-parietal, and visual networks (Figure 3B&C). In addition, the precuneus region was connected to the dorsolateral prefrontal regions where altered grey-matter volume was found in patients with MDD (t(99)=14.86, p = 5.91*10^-27^; Figure 3D).

**Figure 3:**
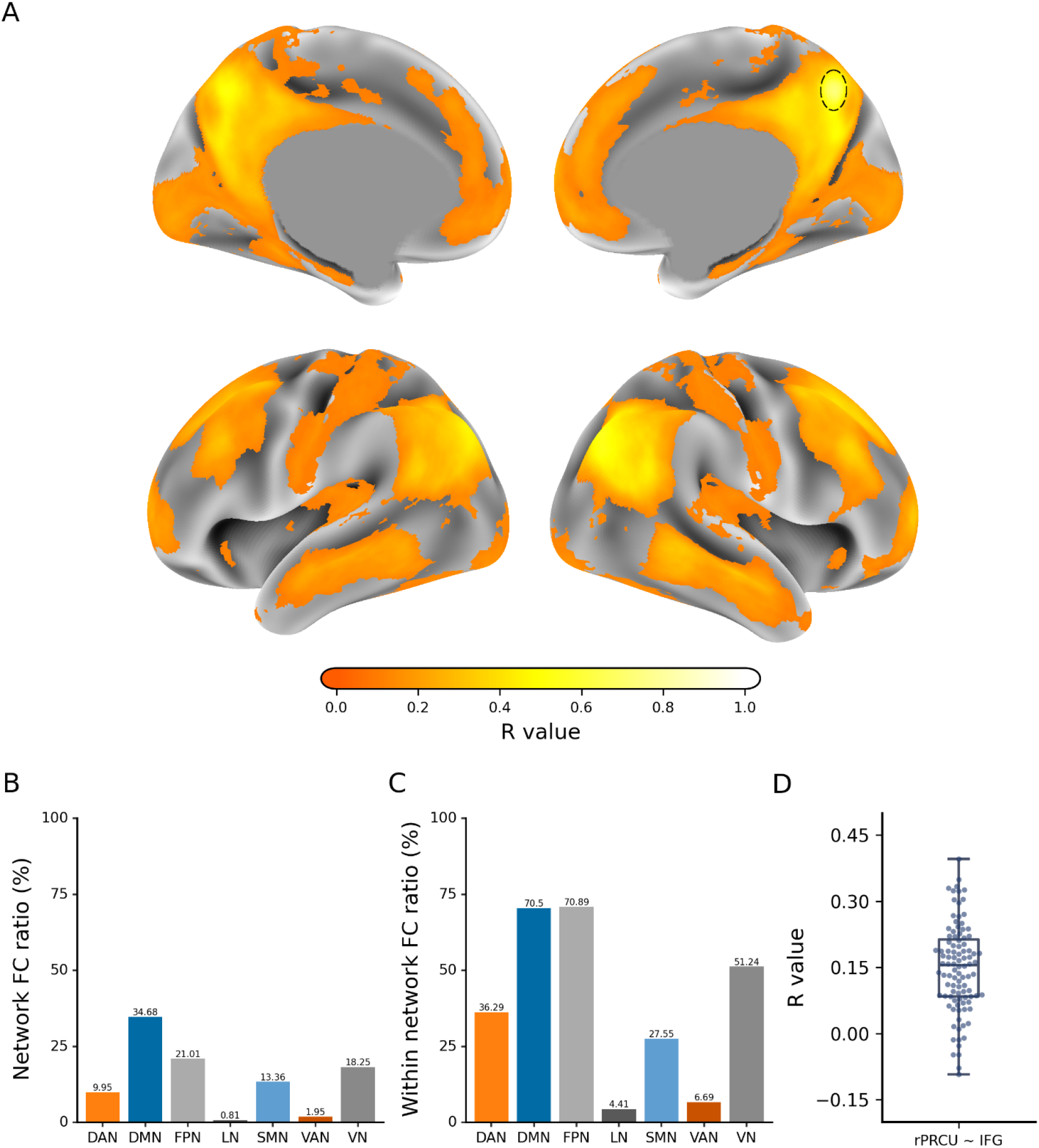
Normative functional connectivity associated with rumination-related region. (A) The precuneus region associated with rumination scores (highlighted on the surface with a black circle) was situated within a normative functional connectivity network. A minimum connectivity strength of r = 0.1 was set. Images are thresholded at P_FWE_< 0.05. (B) The connectivity of the precuneus seed with canonical functional networks (Yeo and Krienen, 2011) was calculated as the proportion of significant vertices found within each network. (C) The proportion of each canonical functional network significantly connected with the precuneus seed is also shown. (D) The rumination-related precuneus seed is functionally connected with the right IFG region where grey-matter volume was found to differ between MDD and control groups. Data points represent connectivity values from HCP dataset participants used to estimate normative functional connectivity. DAN = Dorsal Attention Network; DMN = Default Mode Network; FPN = Fronto-Parietal Network; LN = Limbic Network; SMN = Sensory-Motor Network; VAN = Ventral Attentional Network; VN = Visual Network.

## Discussion

The overall objective of this study was to investigate grey-matter changes in MDD and relate these to rumination. Grey-matter volume was found to be, on average, increased in the bilateral IFG in patients, extending to the dlPFC in the left hemisphere. These changes were not, however, found to correlate with reported rumination. Instead, cortical surface area values in the right precuneus were found to negatively correlate with rumination questionnaire scores. Using normative functional MRI data, it was shown that this precuneus region is functionally connected to the prefrontal areas showing grey-matter changes in MDD.

The dlPFC region identified as having altered grey-matter volume in MDD has been extensively highlighted as being altered in that condition. As well as imaging findings showing structural and functional changes, this region has been targeted for brain stimulation interventions (Chu et al., 2023). Notably though, the majority of previously reported alterations have been of reduced grey-matter in the region, as opposed to the increases reported here (Grieve et al., 2013; Lai, 2013). This difference may be the result of methodological differences. We use a more recently developed method of grey-matter volume estimation that may return different results than alternatives such as voxel-based morphometry (Winkler et al., 2018). Other studies of MDD using this technique have also reported findings that diverge somewhat from prior reports (Bajaj et al., 2019; Zhang et al., 2023). Dissociations between cortical thickness and surface area have also been reported in other conditions, aligning with the picture presented by the unthresholded maps reported here, although more investigation of this question is required (Zhang et al., 2020). At the same time, MDD is a diverse disorder (Drysdale et al., 2017). Some large sample analyses of cortical structure in it have shown inconsistent findings depending on factors such as age, illness of duration, or geographical origin (Schmaal et al., 2017), or only very small group differences (Winter et al., 2022).

This methodological and clinical variability argues for researchers sharing raw data or statistical maps to allow future meta-analytic work that can synthesise large and diverse samples (Costafreda, 2009; Gorgolewski et al., 2016).

Rumination was not found to be correlated with the structural differences in the prefrontal cortex. Instead, higher RRS scores were associated with reduced cortical surface area in the right precuneus. This brain region has been previously linked to episodic memory (Ahmed et al., 2018), and autobiographical memory in particular (Freton et al., 2014; Sreekumar et al., 2018), along with attention to self-related processes (Zhao et al., 2018). These links may point to a role for this region in the focus on self-related thoughts that is seen in rumination. This may be supported by work showing a relationship between increased rumination and altered functional connectivity between the right precuneus and the rest of the brain in both MDD patients and healthy controls (Jacob et al., 2020). Similarly, it has been suggested that there is a dissociation of the precuneus from the rest of the default mode network in patients with MDD (Dutta et al., 2019). Together, these results suggest that the precuneus may be an important node within a wider rumination-related network.

This network-level approach was followed by mapping the normative functional connectivity patterns for the identified rumination-related precuneus region. This showed that the precuneus region is connected to multiple canonical resting-state networks, including the default-mode, fronto-parietal, and visual networks. Importantly, this pattern of connectivity includes the lateral prefrontal regions in which altered grey-matter volume was found in MDD patients. This pattern of functional connectivity follows known white matter connections (Pisner et al., 2019; Tanglay et al., 2022) and is consistent with the status of the precuneus as a “rich-club” hub in the brain (van den Heuvel and Sporns, 2011). Interestingly, a relationship between the structural integrity of the superior longitudinal fasciculus, which links the precuneus with frontal regions, and rumination in MDD has been described (Pisner et al., 2019). More generally, the pattern of connectivity identified corresponds well with prior work highlighting a similar connectivity distribution as a transdiagnostic marker of depression (Siddiqi et al., 2021).

Rumination is a multifaceted phenomenon, and so is likely to be associated with diverse brain processes (Zhou et al., 2020). One may speculate that the functional networks identified in the normative connectivity analysis reported here form an integrative system that can be disrupted by structural alterations in pathological states. This disruption may move the overall of the state from one that supports healthy introspection to one that produces maladaptive rumination. Of the subnetworks highlighted in Figure 3B, the default-mode and fronto-parietal networks have been extensively studied in MDD, with functional and structural alterations reported in areas of each (Kim et al., 2023; Menon, 2011; Sheline et al., 2010).

These networks are associated with rumination-relevant processes such as sustained attention, autobiographical memory, and focus on internal thoughts. A connection between sensory network properties and MDD has also been reported (Furey et al., 2013; Zhang et al., 2023), including changes to structure, function, and biochemical properties (Song et al., 2021; Wu et al., 2023). Notably, the visual system has been connected to rumination in some studies (Burkhouse et al., 2017; Kim et al., 2023; Piguet et al., 2014), where activation may reflect memory-related processes (Dijkstra et al., 2019).

### Limitations

Our study has various limitations. The patient group was mixed in terms of illness onset and duration times. Further, we did not have information about potential depression subtypes within the group. These factors may have influenced the results obtained (Buch and Liston, 2021; Drysdale et al., 2017). Similarly, the patient group was primarily medicated and so effects of antidepressant medication on brain measures cannot be excluded. This may be particularly relevant for measures of grey-matter, where there is some evidence that antidepressant medication can lead to increased cortical thickness (Nemati and Abdallah, 2020). Finally, the sample used was not large, which may influence the statistical results. Recognising this, we have shared raw statistical maps to facilitate cumulative science through future data synthesis.

### Conclusions

This study aimed to identify structural changes in MDD and connect these to reported rumination. It then sought to situate rumination-related regions within a wider network context through normative functional connectivity analysis. We showed that a region in the precuneus was associated with rumination across all participants but that the structure of this region did not differ between patients and controls. Instead, a region in the dlPFC differed in grey-matter volume between groups. This region was situated within the functional connectivity network of the precuneus. Together these results suggest that rumination in depression can be understood as a network-level phenomenon, with structural alterations in certain regions potentially modulating activity properties in others.

## Data Availability

All data produced in the present study are available upon reasonable request to the authors.

## Acknowledgments

The authors thank all participants for their time and effort. They are also grateful to Hsin-Yi Wang and Ching Lin for their help with patient recruitment and data collection, and to the MRI technical staff. This work was supported by funding from the Taiwan National Science and Technology Council to TJL (105-2632-H038-001-MY3), TYH (111-2410-H-038-009-MY2) and NWD (110-2628-H-038-001-MY4). This work was also supported by the Taiwan Ministry of Education Higher Education Sprout Project. The authors report no conflicts of interest.

## Author contributions

PZC: Methodology, Formal analysis, Investigation, Writing - Original Draft, Visualisation;

HCL: Conceptualisation, Investigation;

TJL: Conceptualisation, Funding acquisition;

TYH: Conceptualisation, Methodology, Writing - Original Draft, Supervision; NWD: Conceptualisation, Methodology, Writing - Original Draft, Supervision;

## Data availability

Available upon request.

## Supplementary materials

**Supplementary figure 1:**
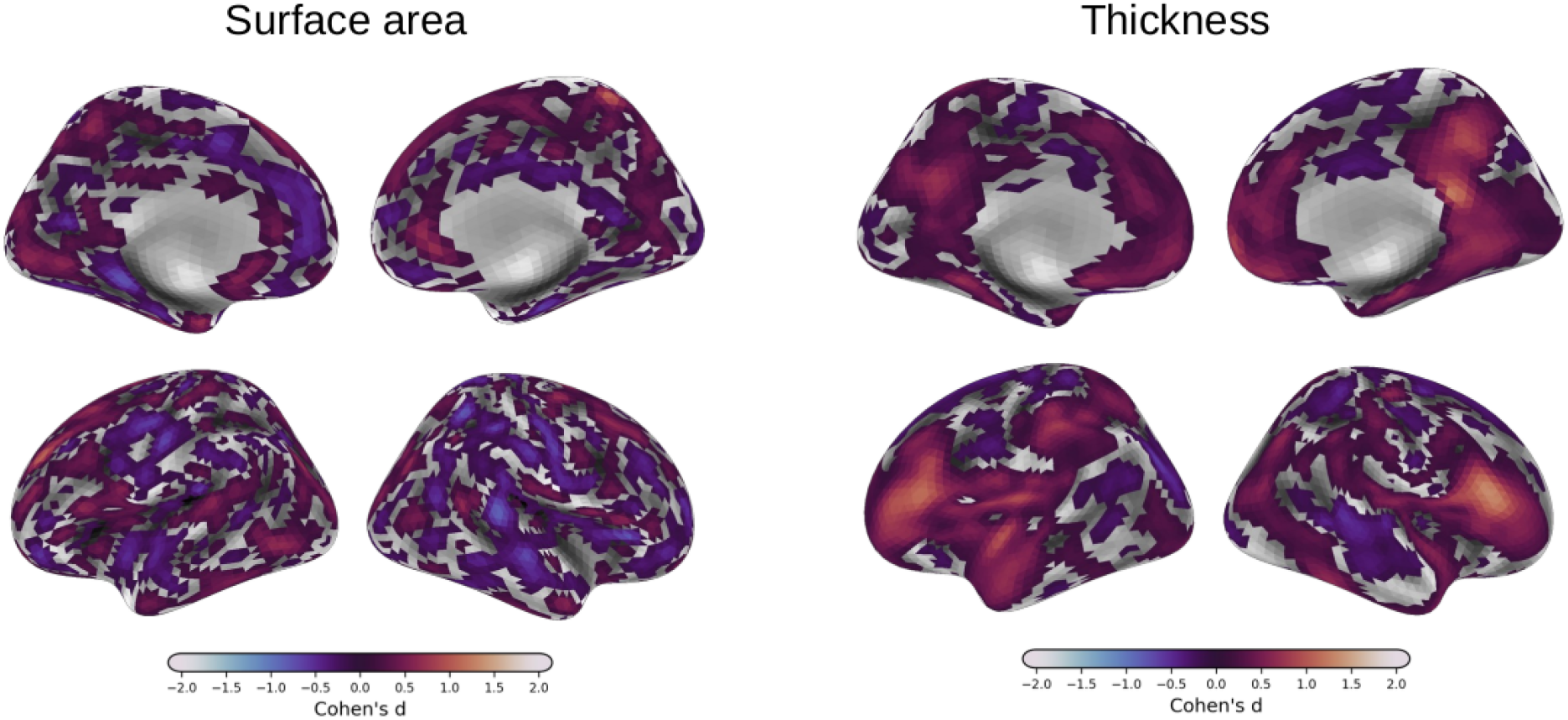
Unthresholded effect-size maps for group comparison of cortical surface area (left) and cortical thickness (right). These metrics are analysed in conjunction for the reported grey-matter volume values.

